# Variation in First-line Type 2 Diabetes Treatment due to eGFR and Provider Preferences: A Novel Statistical Analysis

**DOI:** 10.1101/2024.09.19.24313155

**Authors:** Christina X Ji, Saul Blecker, Michael Oberst, Ming-Chieh Shih, Leora I Horwitz, David Sontag

## Abstract

**Introduction:** The decision between metformin and a DPP-4 inhibitor or sulfonylurea for first-line type 2 diabetes treatment relies on many factors, including estimated glomerular filtration rate (eGFR), history of heart failure, age, sex, and even provider preferences. This study evaluates variation in this treatment decision across two factors: eGFR and provider preferences.

**Research Design and Methods:** Using health insurance claims data, we defined a cohort based on observation prior to first-line treatment, availability of eGFR results, and no type 1 or gestational diabetes (n=10,643). We performed a chi-squared test to verify the association between eGFR and treatment. The cohort was then restricted to providers with at least 10 patients (n=2,271 patients). We conducted a novel statistical analysis to assess variation across providers. We fitted two models to predict treatment—one using only patient characteristics (age, eGFR, sex, history of heart failure, and treatment date) and another using both patient characteristics and provider-specific random effects. With these models, we performed a generalized likelihood ratio test (GLRT) to assess whether including provider-specific random effects improved fit.

**Results:** The chi-squared test confirmed significant association between treatment and eGFR (p < 0.0001). The GLRT in our novel statistical analysis found significant variation existed across providers even after accounting for patient characteristics (p < 0.0001). Visualizations of the observed treatment decisions and treatment policy models show that most of this variation across providers occurred at low eGFR levels, where the level of kidney damage at which metformin should be contraindicated is unclear.

**Conclusions:** While some variation in first-line type 2 diabetes treatment was associated with eGFR, some variation may be due to provider preferences that cannot be explained by treatment guidelines. Further studies can elucidate whether such variation across providers is appropriate. Our approach can be applied to other treatment decisions to improve diabetes management.

**Key Messages:** *What is already known on this topic:* Guidelines for first-line type 2 diabetes treatments recommend metformin unless there are contraindications, such as kidney damage indicated by low estimated glomerular filtration rate (eGFR).

*What this study adds:* This study uses a health insurance claims dataset to verify that first-line treatment is significantly associated with eGFR levels. Then, we propose a novel statistical analysis to assess whether significant variation exists across providers even after accounting for patient age, eGFR, sex, history of heart failure, and treatment date. By fitting two random effects models—one with only patient characteristics and one that also utilizes provider-specific random effects—and comparing the likelihoods of the observed treatment decisions under the two models, we find that the treatment decisions can be explained significantly better when accounting for differences among providers in treatment preferences and eGFR considerations.

*How this study might affect research, practice, or policy:* Our results suggest future studies about whether the significant variation across providers found in our analysis is appropriate may help improve first-line type 2 diabetes treatment decisions, and our novel statistical approach can be applied to evaluate variation across providers throughout the diabetes management process.

## 1. Introduction

Around 1.2 million Americans are diagnosed with type 2 diabetes every year.^1^ Because diabetes can lead to organ damage and other health problems, managing blood glucose levels with lifestyle changes and medication is critical.^2^ This study focuses on the first type 2 diabetes medication patients are prescribed. Specifically, we aim to understand whether variation in this decision can be attributed to patient characteristics and provider preferences.

Variation in treatment decisions due to patient characteristics is supported by treatment guidelines. For instance, the American Diabetes Association (ADA) recommends metformin as the initial treatment for type 2 diabetes unless “there are contraindications”.^2^ Metformin is effective at improving glycemic control and promoting weight loss without increasing risk of hypoglycemia.^3^ However, one contraindication is kidney damage. ADA guidelines currently suggest that metformin is safe in chronic kidney disease (CKD) so long as estimated glomerular filtration rate (eGFR) is at least 30 mL/min/1.73m^2^.^2^ In this study, we first verify that metformin usage decreases with evidence of kidney damage using a health insurance claims dataset.

Variation in treatment decisions due to provider preferences reflects a deviation from guidelines in actual practice. This variation may be due to incomplete or unclear guidelines, barriers to adoption, different practice styles of individual providers and health systems, or other factors that influence patient-provider interactions.^4–5^ For an example of how these factors may come into play, we can again examine the relationship between first-line type 2 diabetes treatment prescriptions and kidney damage. For patients with mild renal impairment, defined as eGFR levels between 30 and 60, metformin only started being recommended in 2016 after several large-scale cohort studies found that metformin did not increase the risk of lactic acidosis in these patients.^6–8^ Because there is uncertainty around the specific eGFR threshold at which the risk of metformin in CKD outweighs the benefits in diabetes, the decision to prescribe an alternative to metformin falls on the individual provider. Other factors may also lead to different treatment preferences among providers.^9^ In this study, we conduct a novel statistical analysis to ascertain whether there is significant variation in first-line type 2 diabetes treatment decisions across providers after accounting for patient characteristics. Past works with similar approaches have examined whether the choice of physician has an impact on cancer treatment and hospice enrollment.^10–12^

Our approach captures decision-making processes that may not be covered by or differ from guidelines. Our method also accounts for differences in the patient populations seen by each provider. For instance, a provider who takes on more challenging cases by seeing patients who have more co-morbidities or are more likely to have contraindications should not be unduly flagged as an outlier. By applying these approaches, this work establishes the existence of variation across providers that cannot be explained by treatment guidelines and relevant patient characteristics. We hope this work will encourage future studies that examine how this variation affects patient outcomes and, if needed, how to issue better guidance to reduce harmful variation.

## 2. Research Design and Methods

### 2.1 Outcome Definition

The outcome of interest was a binary variable for whether the patient was prescribed metformin versus a sulfonylurea or a dipeptidyl peptidase-4 (DPP-4) inhibitor. Although glucagon-like peptide-1 receptor agonists (GLP-1 RA) and sodium glucose cotransporter 2 inhibitors (SGLT2i) became more widely recommended after 2021, sulfonylureas and DPP-4 inhibitors were more commonly prescribed as alternatives to metformin between 2012 and 2021, the period from which the dataset used in this study was collected.^2,13–16^

Diabetes drugs were defined as drugs that are descendants of the “drugs used in diabetes” concept in the Observational Medical Outcomes Partnership (OMOP) Common Data Model (CDM) and contain at least one of the following ingredients: metformin, sitagliptin, vildagliptin, saxagliptin, linagliptin, gemigliptin, anagliptin, teneligliptin, acetohexamide, carbutamide, chlorpropamide, glycyclamide, tolcyclamide, metahexamide, tolazamide, tolbutamide, glibenclamide, glyburide, glibornuride, gliclazide, glipizide, gliquidone, glisoxepide, glyclopyramide, or glimepiride.

### 2.2 Patient Characteristics

Our primary exposure was eGFR, which was based on the last measurement obtained in the 6 months prior to the initial medication fill. Estimated glomerular filtration rate (eGFR) measurements were defined by all lab concepts that started with “Glomerular filtration rate/1.73 sq M”. This included both CKD-EPI that was commonly used through 2017 and MDRD that became more predominant starting in 2018. Because some eGFR measurements are race-adjusted, for Black or African American patients, we prioritized measurement concepts that specified “among Blacks” over concepts that did not specify race and only used concepts that specified “among non-Blacks” if no other measurements were available. For patients of other races or without race specified, the priorities of concepts that specified “among Blacks” and “among non-Blacks” were flipped. If multiple measurement values at the same priority level were taken on the most recent measurement date, the maximum value was selected.

Other variables included age, sex, and the presence of a diagnosis for heart failure in the 2 years prior to type 2 diabetes treatment. Heart failure was defined by concepts that are descendants of the SNOMED concept “Heart failure”. These variables were chosen as they may contribute to variation in decision making related to use of metformin.^17–18^ We also included treatment date as a covariate since metformin usage increased over time, particularly when metformin was no longer contraindicated for patients with moderate renal failure in 2016.^6–8^

### 2.3 Cohort Criteria

We performed a retrospective cohort study using insurance claims data from an insurance provider in the northeast United States, spanning from 2012 to 2021. The database includes laboratory test results when the insurance company has a contract with the laboratory center filling the claim.

Patients who received an initial medication fill for metformin, a sulfonylurea, or a DPP-4 inhibitor were included in the cohort. Patients who were prescribed multiple treatment classes on the initial treatment date, did not have exactly one provider associated with the prescription, or did not have an eGFR measurement in the past 6 months were excluded. To focus on type 2 diabetes, we excluded patients who had a type 1 diabetes, gestational diabetes, neonatal diabetes, or pregnancy-related code at any point in their history. To ensure that the prescription is the first diabetes drug a patient receives, the patient must be observed for at least 95% of the 3 years preceding the treatment date. Although this requirement significantly reduced the cohort size, sufficient prior observation was necessary to ensure that the measured outcome was the first type 2 diabetes medication prescription.

For the analysis on provider-based variability, we additionally limited our cohort to patients who were seen by providers who had at least 10 patients who met the inclusion and exclusion criteria. Treatment decisions were attributed to the provider listed on the insurance claim for the medication fill. The flowchart in Figure 1 depicts the cohort criteria and the number of patients after each criterion was applied.

**Figure 1.**
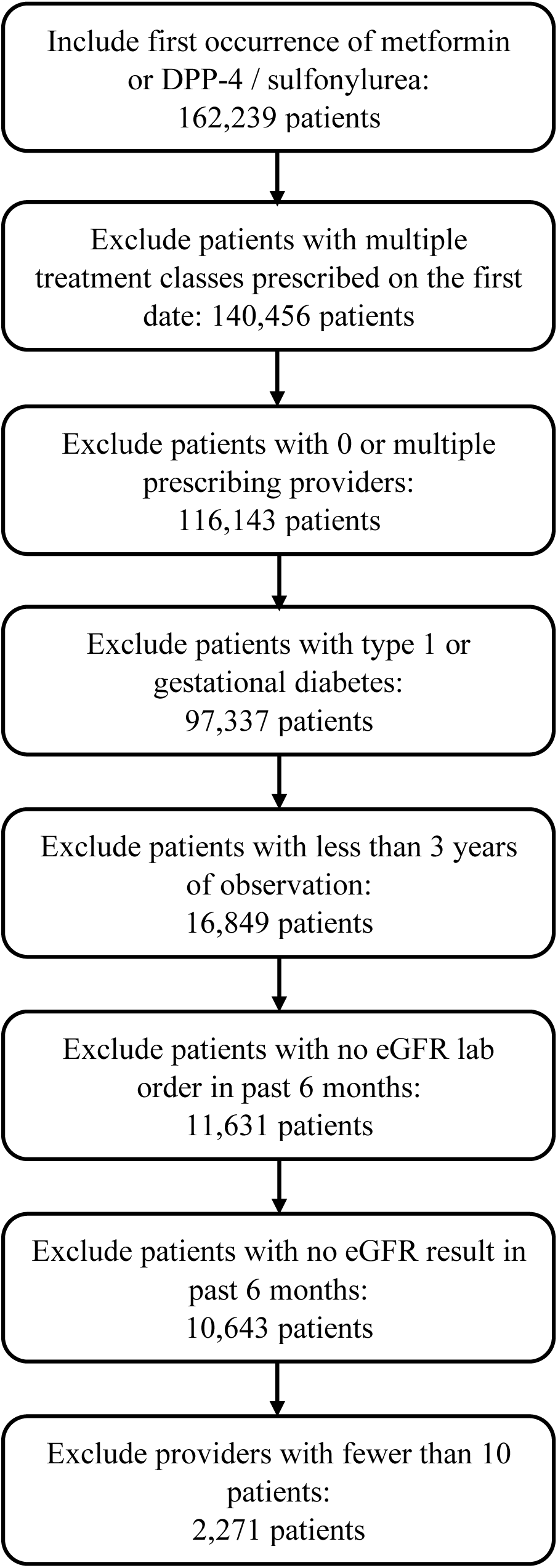
Inclusion-exclusion criteria for cohort.

### 2.4. Statistical Approach to Test for Variation across eGFR Levels

Because kidney damage indicated by low eGFR levels is a known contraindication for metformin, we first verified that the treatment decisions observed in the health insurance claims dataset are aligned with this guideline. We created the following 5 eGFR categories based on the definitions for the different stages of CKD: below 30, 30-44, 45-59, 60-89, and at least 90.^19^ We calculated the number of patients in each eGFR category who were prescribed each medication class. These counts were used in a chi-squared test to determine if there was a difference in rates of metformin use by eGFR category.

### 2.5. A Novel Statistical Approach to Test for Variation across Prescribing Providers

To assess whether providers have different treatment policies with respect to eGFR, we needed to account for how eGFR and other factors affected the likelihood of prescribing metformin. This is important for two reasons: 1) Some providers see more challenging patients.^20^ For example, patients with severe CKD are more likely to seek out specialists than general practitioners for treatment. Modeling patient features accounts for these differences in patient populations across providers. 2) We are interested not just in how providers prescribe the treatments at different rates but in how providers differ in how they take these factors into account when making treatment decisions. Thus, these features need to be included when predicting the likelihood of prescribing metformin.

To account for these features when modeling the treatment decisions, we first found the best model in each of the following two model families: 1) The first model family included generalized linear models with restricted cubic spline features for eGFR, age, and treatment date and binary indicators for heart failure and sex. The cubic spline features allow us to model non-linear relationships and avoid extrapolation issues.^21^ For eGFR, age, and treatment date, we allowed up to 4 knots set at the quantiles suggested by Harrell et al.^21^ 2) The second model family included generalized linear models with the previously mentioned features, random intercepts, and random slopes for the eGFR features. The random intercepts and random slopes reflect differences in how providers account for eGFR and weigh the risks and benefits of each treatment when making decisions. The model can be specified as follows:

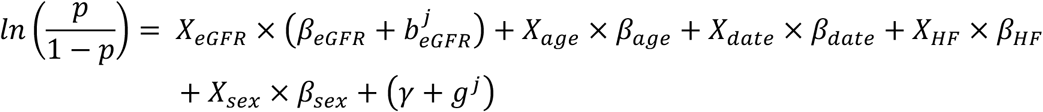

*X*_*eGFR*_, *X*_*age*_, and *X*_*date*_ are vectors containing the feature values and the restricted cubic spline features. *X*_*HF*_ and *X*_*sex*_ are binary indicators for heart failure and sex. The fixed slopes are denoted by *β*, and the random slope for eGFR is denoted by *b^j^_eGFR_* for provider *j*. The fixed intercept is denoted by *γ*, and the random intercept is denoted by *g^j^* for provider *j*. *p* is the probability of prescribing metformin. The random effects follow the structure [*b^j^_eGFR_*, *g^j^*]∼𝒩(0, Σ), where Σ is a parameter that is estimated alongside the fixed slopes and intercept.

We performed a generalized likelihood ratio test (GLRT) to assess whether the data has a significantly higher likelihood under the model with provider-specific random effects.^22–23^ The G-statistic is

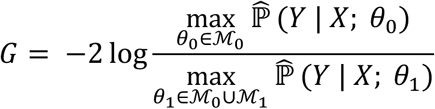

ℳ_0_ is the model family without provider-specific random effects. It is defined by 12 parameters.

ℳ_1_ is the model family with provider-specific random effects. It is defined by 18 parameters. The numerator is the product of the probabilities of the treatment decisions as predicted by the model without provider-specific random effects. The denominator is the product of the probabilities predicted by the model with provider-specific random effects. This G-statistic compares the likelihood of the observed treatment decisions under the two models. A larger G-statistic reflects a better fit from the model with provider-specific random effects. By evaluating the likelihood only at the observed samples, this test focuses on decisions for the types of patients who were actually seen by each provider. The G-statistic follows a *χ*^2^ distribution with 6 degrees of freedom under the null hypothesis.

The method we propose joins an existing body of work on identifying outlying providers, including mixture models of provider effects, hierarchical and Bayesian approaches for modeling random effects, and Markov chain Monte Carlo simulations of normal behavior.^24–28^ Unlike these prior works, our method tests whether variation exists across all providers rather than whether a single provider is an outlier. This is particularly useful when there are few samples per provider. While there may be insufficient power to identify individual providers as outliers, the total number of samples across all providers may provide sufficient power to identify whether variation exists across all providers.

As an additional analysis, we also examined whether any individual provider deviated significantly from the average treatment policy. For this part, we conducted separate GLRTs using only the samples from patients seen by a particular provider. We used the Benjamini-Hochberg procedure to keep the expected false discovery rate at 5%.^29^ In contrast to the simulation approach for identifying outlying providers proposed by Ohlssen et al,^24^ our method does not exaggerate the significance of a small number of decisions from a single provider.

### 2.6. Visualizations

To visualize how the treatment decisions made by each provider vary with eGFR, we first plotted the treatments observed for patients in the dataset against each eGFR value, with the decisions for each provider in a separate row. Then, we plotted the treatment policies learned by the models fit for the GLRT. The model predicts the likelihood of prescribing metformin given eGFR level, age, sex, treatment date, and history of heart failure. We held all features besides eGFR constant, so the policy shown is for a hypothetical patient in their mid-60s to mid-70s with no history of heart failure who is given treatment in mid-2019. These patient characteristics were chosen arbitrarily to fall within the observed population. To create this second plot, we varied the eGFR value from the minimum to the maximum observed (3 to 155) and plotted the predicted likelihood of prescribing metformin in general and for each provider. Unlike the observed decisions in the first plot, the second figure shows the policy a provider may follow at any eGFR value. We also included a histogram depicting the number of similar patients prescribed each medication class in each eGFR range. This histogram shows the range of eGFR values where the model is supported by observed treatment decisions. As such, conclusions should only be drawn about the treatment policy within the eGFR range where there are patients in the histogram.

### 2.7 Data and Resource Availability

The data that support the findings of this study were obtained from Independence Blue Cross and are not publicly available. This research was ruled exempt by MIT’s IRB (protocol E-4025).

Data was extracted using SQL from a postgres database. Models were fit using the glm and glmer packages in R. All other statistical procedures were carried out using Python. All code is publicly available at https://github.com/clinicalml/t2dm_provider_variation_analysis.

## 3. Results

### 3.1. Descriptive Statistics and Variation across eGFR levels

A total of 10,643 patients met the inclusion and exclusion criteria. As shown in Table 1, metformin was prescribed to 83.0% of these patients, and the other 17.0% were prescribed a sulfonylurea or a DPP-4 inhibitor. 6.7% of patients had heart failure in the past 730 days.

**Table 1.** Characteristics of patients and providers in the cohort.

|  | eGFR | Number of<br>patients per<br>provider | Metformin<br>prescription rate<br>per provider |
| --- | --- | --- | --- |
| Minimum | 3 | 10 | 22.7% |
| 25<sup>th</sup> percentile | 70 | 11 | 72.7% |
| Median | 86 | 12 | 85.7% |
| 75<sup>th</sup> percentile | 99 | 14 | 92.9% |
| Maximum | 155 | 41 | 100.0% |
|  | Number of<br>patients | Metformin<br>prescription rate |  |
| Cohort | 10,643 | 83.0% |  |
| Has heart failure | 709 | 64.0% |  |
| No heart failure | 9,934 | 84.4% |  |

Statistics in the eGFR column and the bottom 3 rows are reported for the entire cohort of 10,643 patients. Number of patients per provider and metformin prescription rate per provider are reported for the cohort of 2,271 patients restricted to providers who saw at least 10 patients.

Metformin was prescribed for only 64.0% of patients who had heart failure, compared to 84.4% of patients who did not have heart failure. The mean eGFR level was 84.0, with a standard deviation of 20.5. Most patients have eGFR levels that are indicative of little to no kidney damage as the 25^th^ percentile of eGFR is 70.

As shown in Table 2, both categories of medications were prescribed across the range of eGFR values. However, there was a noticeable and significant increase in the prevalence of metformin prescriptions as eGFR values increased (p<0.0001 from chi-squared test). Only 14.1% of patients with eGFR less than 30 were prescribed metformin. This rate increased to 31.1% among patients with eGFR 30-44 and 66.6% among patients with eGFR 45-59. For patients whose eGFR measurements indicated no sign of kidney damage, metformin prescription rates were 85.3% among patients with eGFR 60-89 and 88.8% among patients with eGFR ≥ 90.

**Table 2.** Number of patients in each eGFR category who were prescribed metformin or DPP-4i / sulfonylurea as their initial diabetes medication.

| eGFR level | Metformin | DPP-4i / Sulfonylurea | Total |
| --- | --- | --- | --- |
| eGFR < 30 | 9 | 55 | 64 |
| $30 \leq \text{eGFR} < 45$ | 92 | 204 | 296 |
| $45 \leq \text{eGFR} < 60$ | 709 | 356 | 1,065 |
| $60 \leq \text{eGFR} < 90$ | 3,851 | 662 | 4,513 |
| $90 \leq \text{eGFR}$ | 4,178 | 527 | 4,705 |
| Total | 8,839 | 1,804 | 10,643 |

### 3.2. Variation across Providers

There were 173 providers who saw at least 10 patients in the dataset. Among these 173 providers, the mean number of patients per provider was 13.1 with a range of 10 to 41. The total number of patients in this restricted cohort was 2,271. As seen in Table 2, the 75^th^ percentile of the number of patients seen per provider was 14. This means most providers saw a small number of patients. The mean per-provider metformin prescription rate was 82.2% with a range from 22.7% to 100.0%. The 25^th^ percentile of the metformin prescription rate per provider was 72.7%, reflecting how most providers prescribe metformin to the majority of their patients.

The maximum likelihood models with and without random effects both had 4 knots each for eGFR, age, and treatment date. The GLRT comparing the data likelihood under the model with random effects and the model without random effects indicated significant variation in metformin usage across providers even when accounting for eGFR, age, sex, prior heart failure, and treatment date (G-statistic 387.3, p<0.0001). This result supports the conclusion that significant variation in treatment decisions may be due to provider preferences that cannot be explained by treatment guidelines.

While there was variation across all providers, we were not able to identify any single provider as an outlier. Because each provider only had a few samples, there is insufficient evidence to conclude with confidence that any provider was an outlier. The smallest p-value for a single provider was 0.059.

Figure 2 illustrates that metformin use increased as eGFR increases. However, there was heterogeneity among providers. Most of this variation among providers occurred at low eGFR values. Some providers still prescribed metformin when eGFR was low, while other providers did not use metformin even when eGFR was high. Providers at the top of the figure prescribed metformin at lower frequencies. Those at the bottom prescribed entirely metformin.

**Figure 2.**
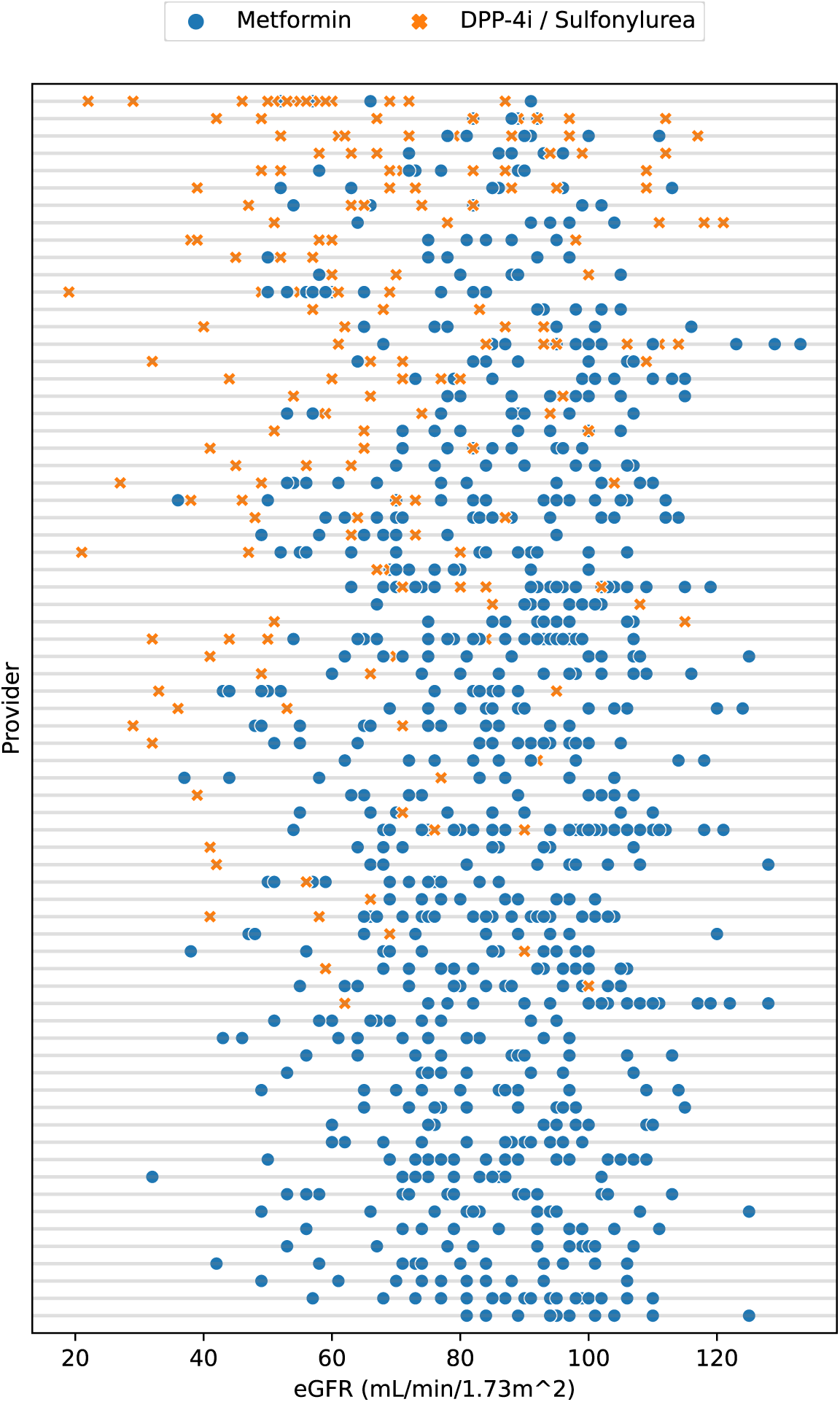
Individual provider choice of initial treatment for diabetes, by most recent eGFR on x-axis. Each row is one provider. Providers ordered by metformin prescription rates. A subset of the 173 providers with at least 10 patients are shown: 10 with lowest rates, 10 with highest rates, and every third in between. A blue dot indicates the provider prescribed metformin to a patient with that eGFR value. An orange X is the corresponding decision to prescribe DPP-4i / sulfonylurea.

Whereas Figure 2 examines the observed treatment decisions in the entire cohort, Figure 3 focuses on the treatment decisions predicted by the models for hypothetical patients with certain characteristics who received treatment on a particular date. When analyzing this figure, we focused on predictions for eGFR values between 40 and 110, where the model is supported by observed treatment decisions. The model may be extrapolating at eGFR values outside this range where no patients are observed. Because the GLRT only evaluates the two models at observed samples, these extrapolated regions have no effect on our test for provider variation.

**Figure 3.**
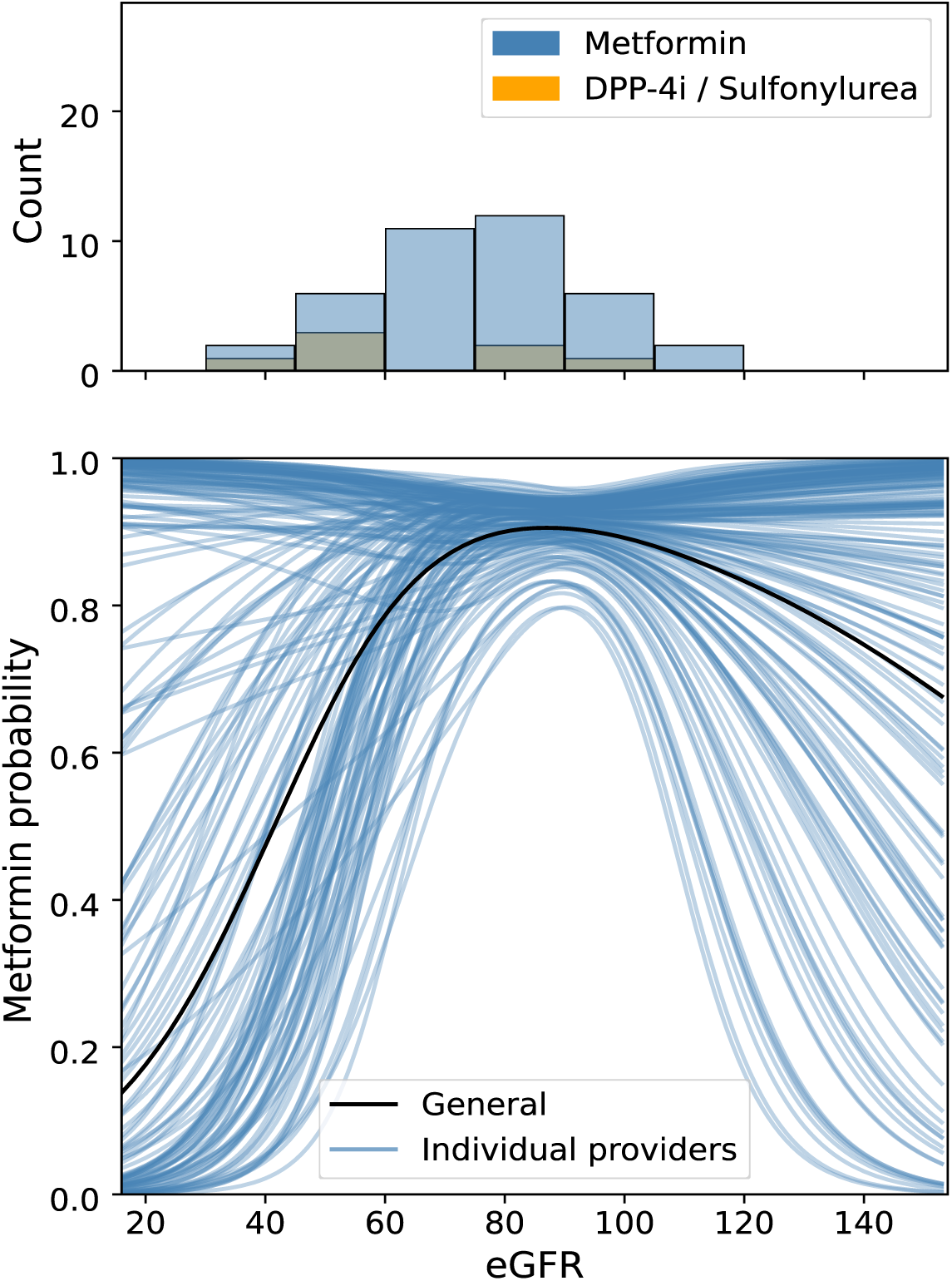
Predicted probability of prescribing metformin versus eGFR for a hypothetical patient in their mid-60s to mid-70s without heart failure whose first-line treatment was initiated in mid-2019. Top: Number of patients who received metformin (blue) and a DPP-4i inhibitor or sulfonylurea (orange) in each eGFR range. Includes patients in their mid-60s to mid-70s of the same sex as the hypothetical patient in the bottom plot who do not have a history of heart failure and received treatment between late 2017 and late 2020. Bottom: Metformin prescription probabilities estimated for the hypothetical patient from models without random effects (black) and with provider-specific random effects (blue).

The black line in Figure 3 depicts the model learned without provider-specific random effects. This model of the general prescribing practice across all providers predicts low metformin prescription probabilities around 40% for small eGFR values. The predicted probabilities increase to around 90% as eGFR levels start to indicate normal kidney function. The blue lines in Figure 3 depict the policies learned for each provider. Not all providers saw patients with low eGFR values, so some of those policies may not be supported. Nonetheless, in general, when eGFR was low, some providers still prescribed metformin with high probability, while other providers were less inclined to give metformin. Similar trends were observed for hypothetical patients with other characteristics that were observed in the cohort.

## 4. Discussion

Using a health insurance claims dataset, we verified that the choice of first-line treatment for type 2 diabetes was associated with eGFR level and demonstrated in a novel statistical analysis that the first-line treatment decision also varied significantly with prescribing provider. The first part of our study verified that observations in our cohort aligned with prior studies.^13–15^ We observed that metformin was prescribed to a large proportion of patients. We also confirmed that metformin was prescribed significantly more frequently to patients with higher eGFR levels, as would be expected based on treatment guidelines. The second part of our study then established a novel empirical result: eGFR level, age, sex, history of heart failure, and treatment date did not fully explain variation in metformin prescription. Significant variation across providers existed in the first-line type 2 diabetes decisions. This suggests that if a patient sees different providers, they may receive different care, be prescribed different treatments, and ultimately have different outcomes.

The degree of provider-level variation is particularly interesting because of its novelty and the potential for interventions to improve patient care. Most prior research in this area relied on interviews and surveys asking providers about their awareness of contraindications to metformin, such as the relatively recent guideline that lowered the eGFR cutoff at which metformin is no longer recommended to 30.^17,30^ These surveys may not always reflect clinical practice.^31^ Other past studies related to variation in clinical practice for type 2 diabetes focused on the association between patient-reported outcomes and process-of-care indicators regarding whether lab measurements and annual examinations were made.^32–35^ These studies did not consider the treatment decision-making process.

To our knowledge, this is the first study that uses an observational dataset to empirically assess how first-line type 2 diabetes treatment varies across providers while accounting for patient characteristics. Using the observational dataset allowed us to draw conclusions about how providers differ in the decisions that were actually made. Observational datasets have been used to study first-line type 2 diabetes treatments in prior works on changes in prescribing patterns over time and cardiovascular risks associated with different treatments.^36–37^ This is the first work, to our knowledge, to leverage observational data to study variation in first-line type 2 diabetes treatment decisions across providers.

Because this study demonstrated that variation exists across providers who prescribe first-line type 2 diabetes treatments, it raises the question of whether this variation needs to be addressed. If the variation is inappropriate, how can the negative effects be mitigated? A non-specific intervention would be to provide brief education about first-line diabetes treatments to providers whose treatment decisions do not align with the guidelines related to eGFR and metformin. More data-driven and individualized interventions such as reviewing provider practice patterns to identify areas for improvement or imposing financial penalties for unwarranted variation would require accurate assessment of the performance of individual clinicians, identification of outlying providers, and good intervention design.^38–40^ For such interventions, there must be sufficient evidence based on decisions for a large number of patients that the provider is indeed making poor treatment decisions to avoid incorrectly penalizing providers.^41–42^ As we saw in our tests for individual providers, there was an insufficient number of patients per provider for us to draw any conclusions about individual providers.

Our study should be interpreted in the context of its limitations. First, this study was limited to a single insurance provider whose beneficiaries are primarily located in the northeast United States. As a result, the findings may not be generalizable. Second, while our models included several patient characteristics, they may not have accounted for all patient-level factors that contribute to treatment decisions. Third, while we chose to evaluate the performance of individual providers, variation may also be studied at the level of provider groups or health care facilities.^35^

The statistical approach in this study is more broadly applicable to other treatment decisions. The models in our method can be built with relevant patient characteristics for those decisions. Identifying variation in treatment practices that cannot be explained by guidelines opens the door to improvements in guidelines and practices for diabetes management.

## Data Availability

The data is not publicly available.

## Acknowledgments

*Personal Thanks:* We would like to thank Rebecca Boiarsky and Justin Lim for setting up the databases used in this study. We would also like to thank James Denyer, Aaron Smith-McLallen, Stephanie Gervasi, and the rest of the data science group at Independence Blue Cross for providing the dataset.

*Funding and Assistance:* This work was supported in part by Independence Blue Cross, Office of Naval Research Award No. N00014-21-1-2807, and the LEAP program from the Ministry of Science and Technology in Taiwan.

## Author Contributions and Guarantor Statement

Christina X Ji: Conceptualization, Data curation, Formal analysis, Investigation, Methodology, Software, Validation, Visualization, Writing – original draft, Writing – review & editing; Saul Blecker: Conceptualization, Validation, Writing – original draft, Writing – review & editing; Michael Oberst: Conceptualization, Data curation, Formal analysis, Investigation, Methodology, Software, Validation, Visualization; Ming-Chieh Shih: Conceptualization, Data curation, Formal analysis, Investigation, Methodology, Software, Visualization; Leora I Horwitz: Conceptualization, Validation, Writing – review & editing; David Sontag: Conceptualization, Funding acquisition, Supervision

## Preprint

This work has been posted on the medrxiv preprint server at https://www.medrxiv.org/content/10.1101/2024.09.19.24313155.

